# Integration of Clinicopathological And Genomic Features To Predict The Risk Stratification of TCGA Lung Adenocarcinoma And Lung Squamous Cell Carcinoma Patients

**DOI:** 10.1101/2022.07.14.22277645

**Authors:** Mehmet Cihan Sakman, Talip Zengin, Tuğba Önal-Süzek

## Abstract

**Background:** Predicting lung adenocarcinoma (LUAD) and Lung Squamous Cell Carcinoma (LUSC) risk cohorts is a crucial step in precision oncology. Currently, clinicians and patients are informed about the patient’s risk group via staging. Recently, several machine learning approaches are reported for the stratification of LUAD and LUSC patients, but there is no study comparatively assessing the integrated modeling of the clinicopathological and genetic data of these two lung cancer types so far.

**Methods:** In our study based on 1026 patients’ clinicopathological and somatically mutated gene features, a prognostic prediction model is implemented to rank the importance of features according to their impact on risk classification.

**Findings:** By integrating the clinicopathological features and somatically mutated genes of patients, we achieved the highest accuracy; %93 for LUAD and %89 for LUSC, respectively. Our second finding is that new prognostic genes such as KEAP1 for LUAD and CSMD3 for LUSC and new clinicopathological factors such as site of resection are significantly associated with the risk stratification and can be integrated into clinical decision making.

**Conclusions:** In current clinical practice, clinicians, and patients are informed about the patient’s risk group only with cancer staging. With the feature set we propose, clinicians and patients can assess the risk group of their patients according to the patient-specific clinical and molecular parameters. Using this machine learning model we are implementing a user-friendly web interface for clinicians and lung cancer patients to predict the risk stratification of individuals and to understand the underlying clinical and molecular mechanisms.

## 1 INTRODUCTION

Lung cancer is the most common type of cancer and the leading cause of death worldwide. The World Health Organization (WHO) reported that lung cancer is the second most frequently diagnosed cancer type, constituted 11.4% of all cancers, and the leading cause of cancer-related deaths (18%) in 2020[1]. In the United States, only 14% of patients who develop lung cancer survive for five years. These mortality rates (>150,000/year) far exceed those of the acquired immunodeficiency syndrome epidemic. However, this survival rate has only slightly increased in the last two decades, and it appears unlikely that marked improvements will occur in the near future[2].

Lung cancer can be divided into two classes: Small Cell Lung Cancer (SCLC) and Non-Small Cell Lung Cancer (NSCLC). Lung Adenocarcinoma (LUAD) and Lung Squamous Cell Carcinoma (LUSC) are two of the three subtypes of Non-Small Cell Lung Cancer (NSCLC) occurring in 85% of lung cancer patients. When the health conditions of LUAD and LUSC cancer patients, which make up most of the lung cancer cases, are detected at an earlier stage, patient risk group based treatment can be applied according to the course of cancer.

Presently, Machine Learning methods are getting integrated into decision making processes at the clinic due to their success in classification and prediction helping medical practitioners[3]. Machine learning provides the solution for decreasing the increasing price of health care and creating improved patient-clinician communication.

In this study, we comparatively evaluated the prediction power of five different machine learning algorithms to identify the risk groups of NSCLC : Support Vector Machine, Logistic Regression, Naive Bayes, Random Forest, and K Neighbors Classifiers using Python sci-kit-learn library.

## 2 BACKGROUND

Among the recent studies in the field of machine learning based lung cancer prediction, several of them used Computed Tomography (CT)[4] whereas other techniques are more specific, and used genomic or phenotypic information for building classification models[5].

In [4] authors used Support Vector Machine on Computed Tomography Images to detect lung malignant tumor cells. Sherafatian and Arjmand instead proposed a Decision Tree-based classifier for lung cancer diagnosis[5]. The proposed learning algorithm applied to miRNA sequencing data and clinical data on 1,068 samples from two lung cancer projects (LUAD and LUSC) in TCGA. The class imbalance issue in training and testing datasets was addressed separately using the Synthetic Minority Oversampling Technique (SMOTE) and the Decision Trees algorithms were performed using the RPART package. In our study we analyzed the prediction power of several machine learning algorithms using the previously unexplored combined features of the clinicopathological properties and somatically mutated driver genes.

## 3 MATERIALS AND METHODS

## 3.1 Data Collection

The genetic data and the corresponding clinical information for LUAD and LUSC were downloaded from the publicly available The Cancer Genome Atlas (TCGA) database. TCGA contains the largest known publicly available cancer patient data, including data from 11,000 patients from 33 different cancer types (https://www.cancer.gov/tcga). In our project, we downloaded the 522 LUAD cancer patients and 504 LUSC cancer patients with corresponding clinical and genetic information. After filtering out the missing values a total of 504 LUAD cancer patients with 51 of them live more than five years and 494 LUSC cancer patients with 83 of them live more than five years.

## 3.2 Preprocessing

A common problem encountered while training machine learning with biological data is its imbalanced and complicated nature due to missing values among data quality limitations. Most of the data in the biomedical domain is not smooth. Therefore, to recoup and preserve valuable biomedical data we applied several pre-processing strategies.

To deal with the high ratio of the missing values, features with less than %80 data content are removed from the training and test datasets. Submitter_id, diagnosis_id, exposure_id, demographic_id, treatment_id, and bcr_patient_barcode columns are random numbers thus do not contain information and are removed from the dataset. Columns that contain duplicate columns such as year_of_birth, state, up_dated_datetime, tissue_or_organ_of_origin are also removed. We also discarded the year_of_birth column as we can find the patient’s age from the days_to_birth(in days) column. In the state feature, all the values are the same as ’released’; therefore, we dropped it. For the columns that contain duplicate value such as (tissue_or_organ_of_origin / site_of_resection_or_biopsy) we dropped one of them.

Data were partitioned into training (80%) and testing (20%) datasets using the sci-kit-learn’s model_selection package and all subsequent exploratory data analysis and model training was performed only on the training dataset.

## 3.3 Missing Value Imputation

The most common two strategies to cope with the missing value problem are dropping the null values and filling out the missing values with the mean of the feature. Although the effectiveness of these approaches is questionable, we experimented with filling out the missing values in train and test data with the mean of the training data.

However, there is an exceptional condition in our dataset that should not be filled with imputation methods. Days to death can exceed the days to the last follow-up, and days to death is not available for patients still living. Therefore, we have experimented with three different methods to deal with that problem:

a. Drop the days_to_last_follow_up and assign 0 for all alive patients in days_to_death, then drop if there is a dead patient with a days_to_death value is 0. As we can see, we have found a 0.63 correlation with vital status, but we could not find any correlation with the patient’s risk.
b. Assume that all the patients will live until 90 years old and fill the NaN values for alive patients assuming that they will live until their 90s. As we can see, we have found a 0.83 correlation with vital status, but we could not find any correlation with the patient’s risk.
c. We transferred the alive patient information into the days_to_death column and dropped the days_to_last_follow_up column. As we can see, we could not find any correlation for days to death neither between vital status nor risk of the patient.

After computing the correlation coefficients of the features for these three strategies (Figure 1), alive patients’ days_to_death values are filled with the days_to_last_follow_up feature.

**Figure 1:**
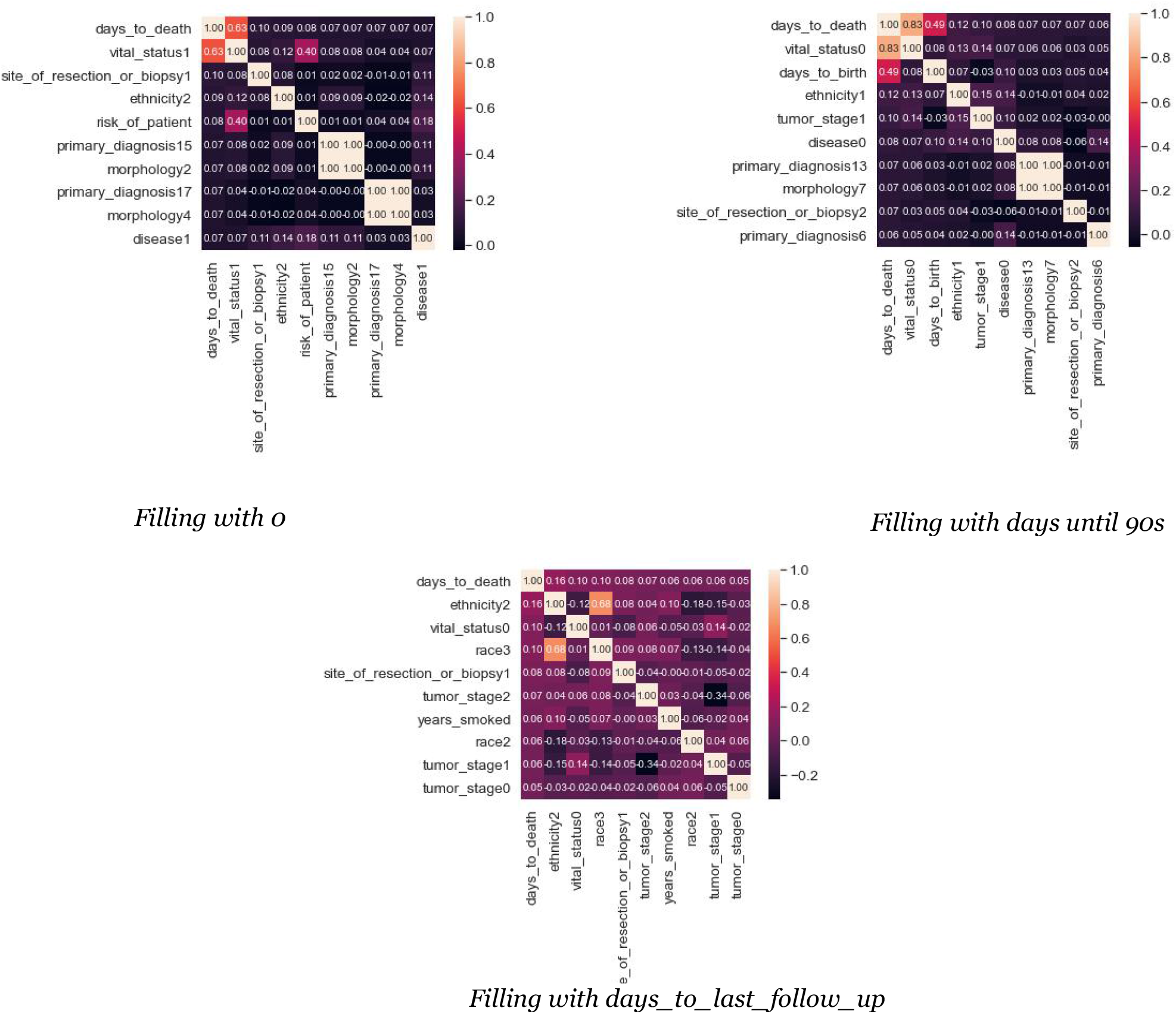
Correlation coefficients of three strategies for assigning missing values of days_to_death column

## 3.4 Numeric Columns Imputation

The missing values in the columns (*age_at_diagnosis, days_to_birth, years_smoked, cigarattes_per_day*) of both LUAD and LUSC training and test datasets are filled with the mean values of the training data using the *SimpleImputer* library in scikit-learn. The column with the most missing value was the *years_smoked* for both LUAD and LUSC. The ratio of missing values in the *years_smoked* feature is 62.83% in LUAD and 55.55% in LUSC.

### 3.5 Categorical Columns Imputation

There are two critical categorical columns in the dataset; race and ethnicity. Nevertheless, 12.8% race and 24.1% ethnicity values are missing for LUAD, and on the other hand, 22.4% race and 35.1% ethnicity values are missing for the LUSC dataset. These two features may directly affect the remaining lifetime, and the authors did not want to impute these missing values. Therefore, missing values for both race and ethnicity are kept as unknown values.

Categorical variables contain label values rather than numeric values such as the “color” variable with the values: “red”, “green”, and “blue”. Each value represents a different category. The main problem with categorical data is that many machine learning algorithms cannot operate on label data directly. Most machine learning algorithms require all input variables and output variables to be numeric. Therefore, we should encode our categorical data into numeric data. To achieve that, we utilized the scikit-learn library’s preprocessing. For the first step, we applied LabelEncoder to categorize each unique value into an integer value. After that, OneHotEncoding is applied to the integer representation. The integer encoded variable is removed, and a new binary variable is added for each unique integer value. This strategy is applied to all our categorical values as depicted in Figure 2.

**Figure 2:**
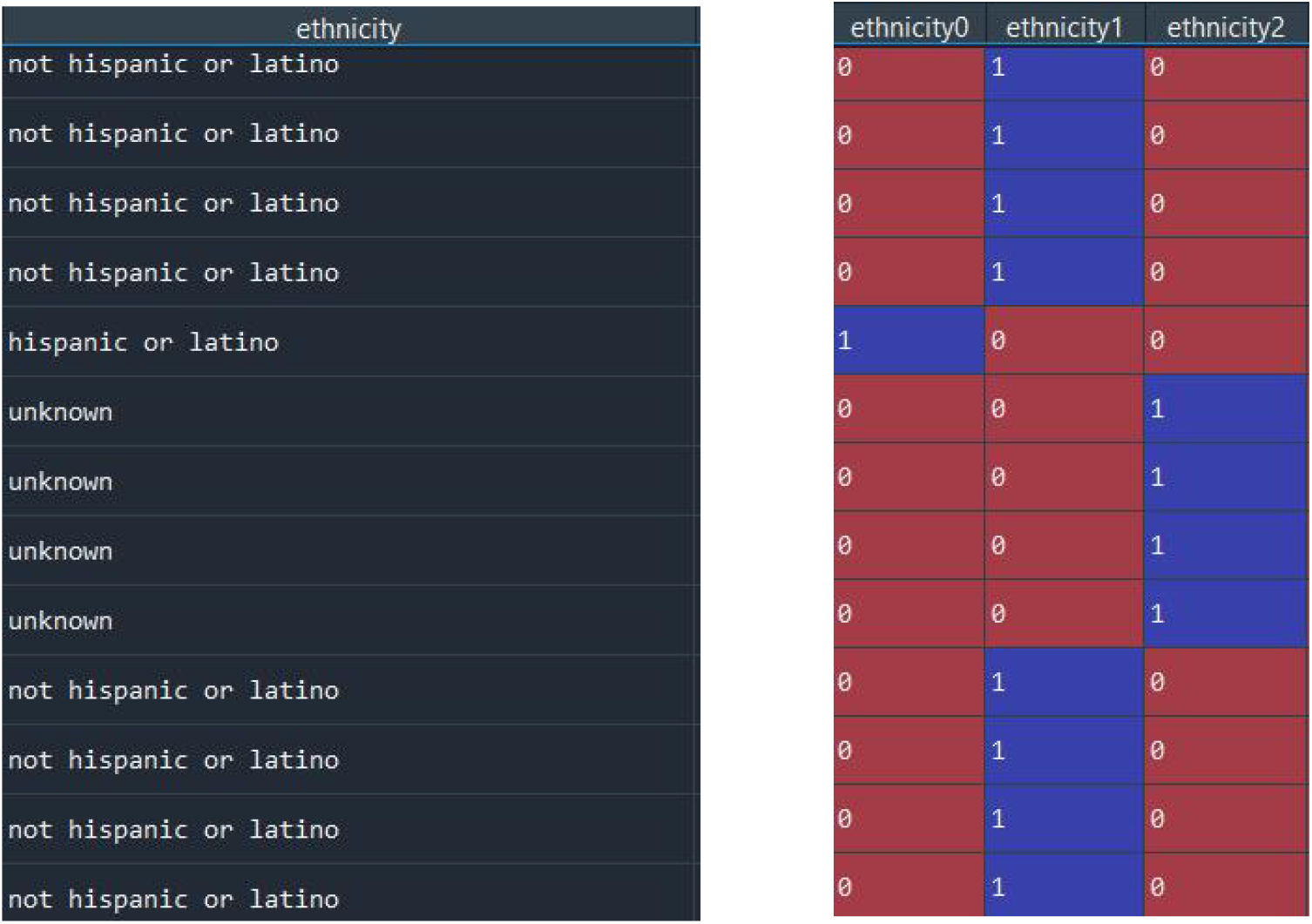
OneHotEncoding of categorical values

### 3.6 Balancing the Imbalanced Classes

For the LUAD and LUSC datasets, the number of patients surviving more than 5 years is approximately 9 times smaller than the opposite. Oversampling and Undersampling lead to similar performances provided that the sampling is correctly implemented on the training and testing folds separately [6]. For the TCGA dataset specifically, several previous studies applied. SMOTE for the TCGA training set [5,8]. Therefore, to cope with the class imbalance problem between the number of positives (i.e. patient survives longer than 5 years) and negatives we over-sampled both of the TCGA LUAD and TCGA LUSC training and testing patient set via Synthetic Minority Oversampling Technique (SMOTE). The class distributions before and after applying the SMOTE algorithm are presented in Figure 3.

**Figure 3:**
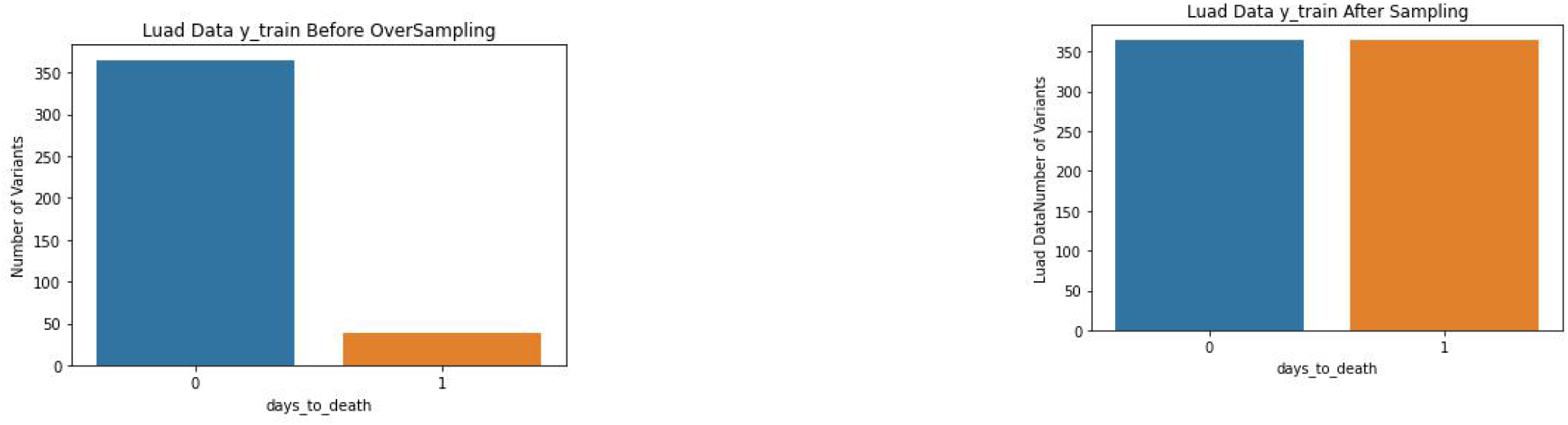
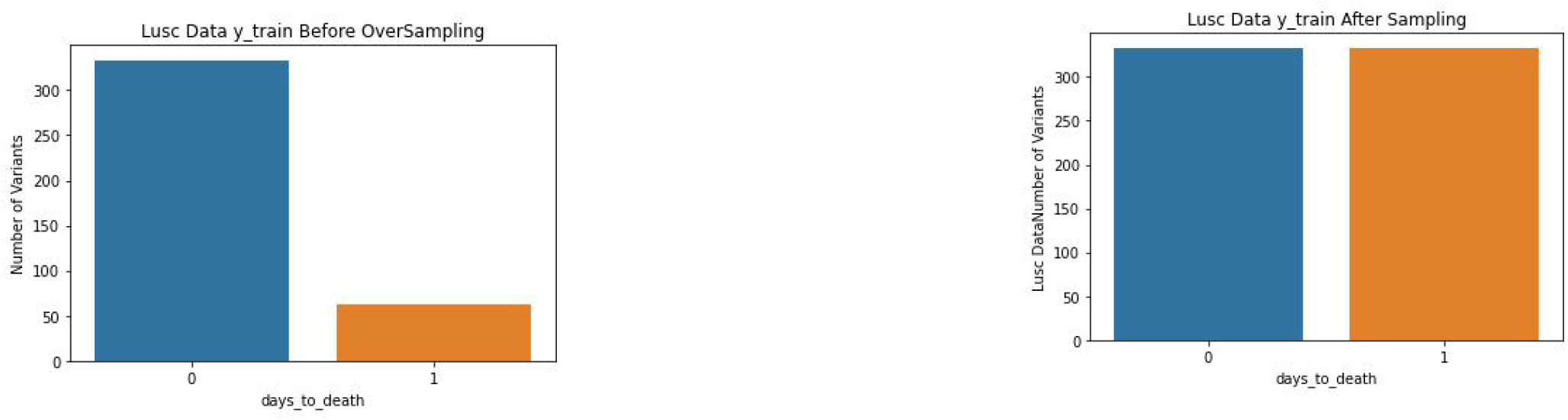
The Class distribution before and after applying the SMOTE algorithm

### 3.7 Creation of the Machine Learning Models

After the data preparation, five different classification algorithms (Logistic Regression, Random Forest Classifier, Naïve Bayes, SVC, and K-Neighbors Classifier) were applied to the LUAD and LUSC datasets. Subsequently, to evaluate the performance of the learning algorithms, the area under the receiver operating characteristics (ROC) curves (AUC) were plotted and calculated (Figure 4,5).

**Figure 4:**
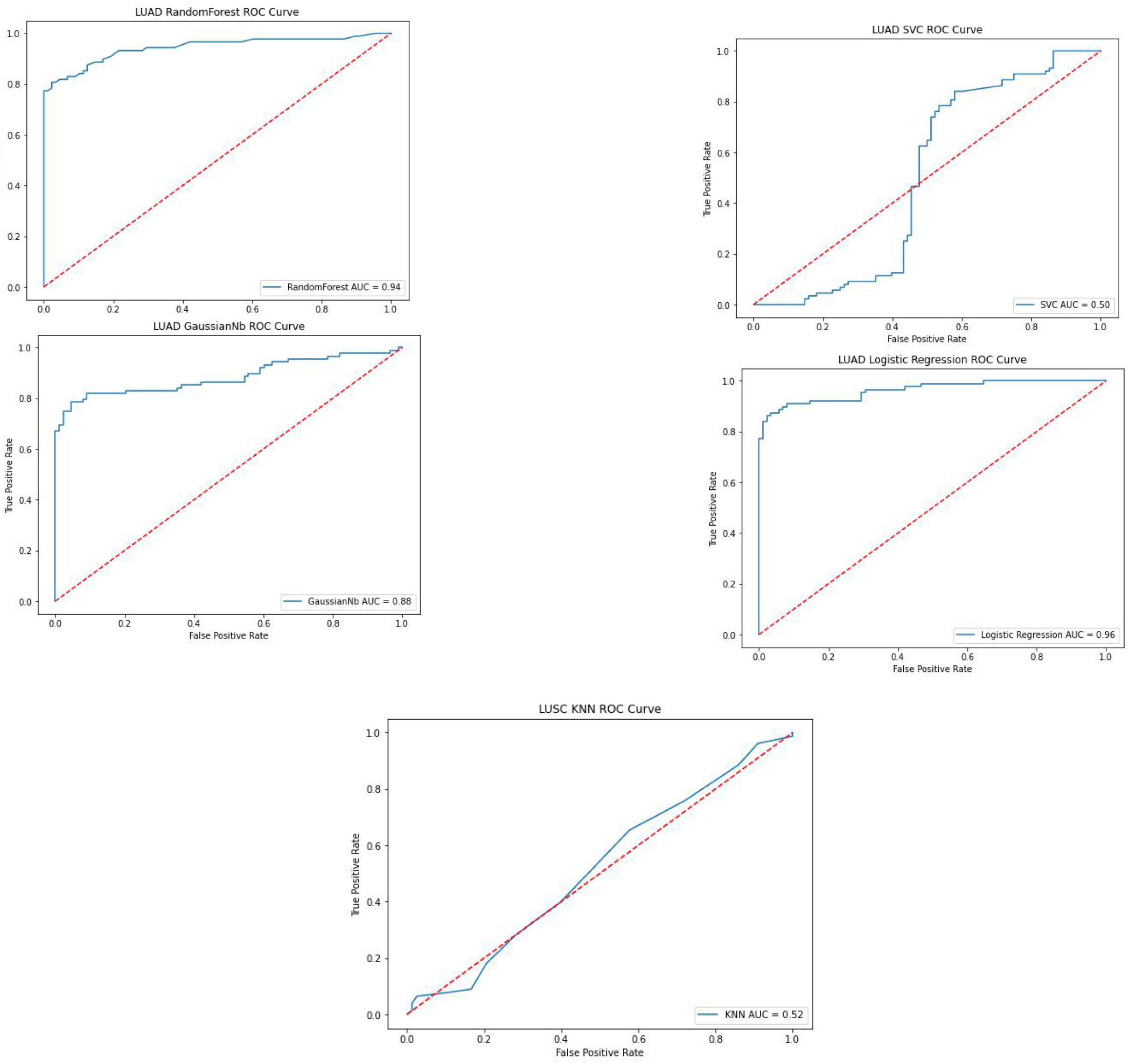
AUC of 5 Algorithms for LUAD

**Figure 5:**
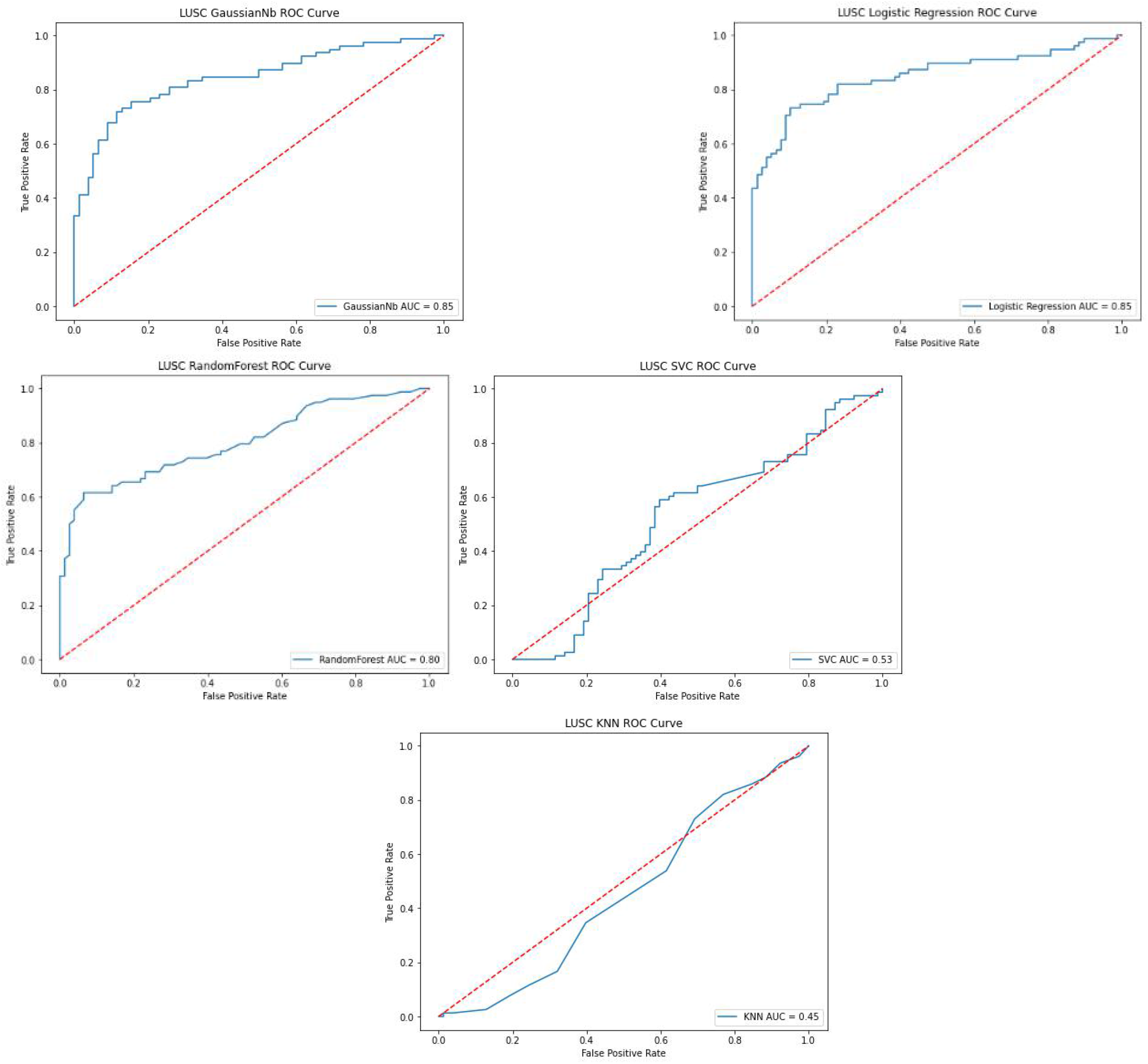
AUC of 5 Algorithms for LUSC

The AUC has been used for decades in the medical area for model selection. It is also commonly used in machine learning algorithms to evaluate the performance of the algorithms [11,12]. It is defined as a plot of a model’s true positive rate as the y-coordinate versus its false-positive rate as the x coordinate, under all possible score thresholds. In order to support the AUC metrics, the F1-score, precision, and recall are calculated and shown in Table 1 and Table 2.

**Table 1:** Comparison of the Precision, Recall, F1-score of 5 algorithms for LUAD

| Metrics of LUAD Algorithm Models |  |  |  |  |
| --- | --- | --- | --- | --- |
| Models | Precision | Recall | F1-Score | Accuracy |
| Logistic Regression | 0.90 | 0.89 | 0.89 | **0.89** |
| Random Forest | 0.82 | 0.81 | 0.81 | 0.81 |
| Naive Bayes | 0.72 | 0.65 | 0.62 | 0.71 |
| SVC | 0.53 | 0.52 | 0.45 | 0.53 |
| KNN | 0.62 | 0.62 | 0.62 | 0.62 |

**Table 2:** Comparison of the Precision, Recall, F1-score of 5 algorithms for LUSC

| Metrics of LUSC Algorithm Models |  |  |  |  |
| --- | --- | --- | --- | --- |
| Models | Precision | Recall | F1-Score | Accuracy |
| Logistic Regression | 0.75 | 0.74 | 0.73 | **0.74** |
| Random Forest | 0.71 | 0.69 | 0.68 | 0.68 |
| Naive Bayes | 0.70 | 0.62 | 0.58 | 0.71 |
| SVC | 0.56 | 0.53 | 0.48 | 0.56 |
| KNN | 0.53 | 0.53 | 0.52 | 0.53 |

### 3.8 Hyperparameter Tuning

Among the five different classification algorithms, the top two best scoring algorithms were found as Random Forest and Logistic Regression, thus hyperparameter tuning is applied to Random Forest and Logistic Regression based models. For hyperparameter tuning of these two algorithms, we implemented a 5-fold cross-validation where we first split the training set into 5 folds and then applied random oversampling on 4 folds which were used for training the classification model and then documented the model performance metrics on the remaining 1-fold using the GridSearchCV in scikit-learn.

## 4 RESULTS

The overall statistics of LUAD and LUSC datasets were summarized separately in Table 4. There are 522 (242 male, 280 female) patients for LUAD, 504 (373 male, 131 female) patients for LUSC. In LUAD, 188 patients and in LUSC 220 patients died during the TCGA project. These patients are divided into four different tumor stages, and they have different smoking habits for different ages. We applied GridSearchCV in scikit-learn to find the best parameters for the models. GridSearchCV is preferably used to tune hyperparameters compared to other tuning algorithms [13].

After applying the scikit-learn’s GridSearchCV (*scoring=’f1’, cv=5*) algorithm to Random Forest the best parameters found as *max_depth=8, max_features=log2, min_samples_leaf=3, n_estimators=50* in LUAD with 0.93 f1-score, and *max_depth=8, max_features=’auto’, min_samples_leaf=3, n_estimators=200* in LUSC with 0.87 f1-score. On the other hand, for the Logistic Regression; GridSearchCV tuned the parameters such as *C=16*.*77, penalty=l2, solver=newton-cg* in LUAD with 0.92 f1-score and C*=109*.*85, penalty=l1, solver=liblinear* with 0.85 f1-score in LUSC (Table 3).

**Table 3:** Comparison of performance metrics for Random Forest and Logistic Regression with and without hyperparameter tuning

| Model | Data | GridSearchCV | Precision | Recall | F1-Score | Accuracy |
| --- | --- | --- | --- | --- | --- | --- |
| Logistic Regression | LUAD | No/Yes | 0.92/**0.94** | 0.92/**0.93** | 0.92/**0.93** | %91.7/%**93.1** |
| Random Forest | LUAD | No/Yes | 0.88/**0.90** | 0.88/**0.90** | 0.87/**0.90** | %87.9/%**89.7** |
| Logistic Regression | LUSC | No/Yes | 0.77/**0.82** | 0.74/**0.78** | 0.74/**0.77** | %74.3/%**77.5** |
| Random Forest | LUSC | No/Yes | 0.72/**0.77** | 0.68/**0.71** | 0.66/**0.69** | %66.6/%**70.5** |

## 4.1 K-Fold Cross Validation

K-fold cross-validation is an approach that splits the dataset into k-fold and shuffles the training and test sets k times to assess how the results of the analysis will generalize to an independent data set[14,15]. We applied 5-fold cross-validation with Logistic Regression and Random Forest models to improve the reliability of these algorithms. During this process, we constructed the Logistic Regression and Random Forest models with hyperparameters obtained from GridSearchCV. Random Forest and Logistic Regression models’ results for each five-folds can be observed in Table 5 and Table 6 with a mean and standard deviation of folds.

**Table 4:** Clinical properties of the LUAD and LUSC patients

| LUAD Category | Number | LUSC Category | Number |
| --- | --- | --- | --- |
| Age at diagnosis (median; range) | 67 (33-89) | Age at diagnosis (median; range) | 68 (39-90) |
| Gender |  | Gender |  |
| Female | 280 | Female | 131 |
| Male | 242 | Male | 373 |
| Number of cigarettes per day (mean; range) | 2 (0-9) | Number of cigarettes per day (mean; range) | 3 (0-13) |
| Number of years smoked (mean; range) | 32 (2-64) | Number of years smoked (mean; range) | 40 (8-63) |
| *Tumor Stage* |  | *Tumor Stage* |  |
| I | 279 | I | 245 |
| II | 124 | II | 163 |
| III | 85 | III | 85 |
| IV | 26 | IV | 7 |
| NA | 8 | NA | 4 |
| *Vital Status* |  | *Vital Status* |  |
| Alive | 334 | Alive | 284 |
| Dead | 188 | Dead | 220 |
| *Ethnicity* |  | *Ethnicity* |  |
| Hispanic or Latino | 7 | Hispanic or Latino | 8 |
| Not Hispanic or Latino | 389 | Not Hispanic or Latino | 319 |
| NA | 126 | NA | 177 |

**Table 5:**
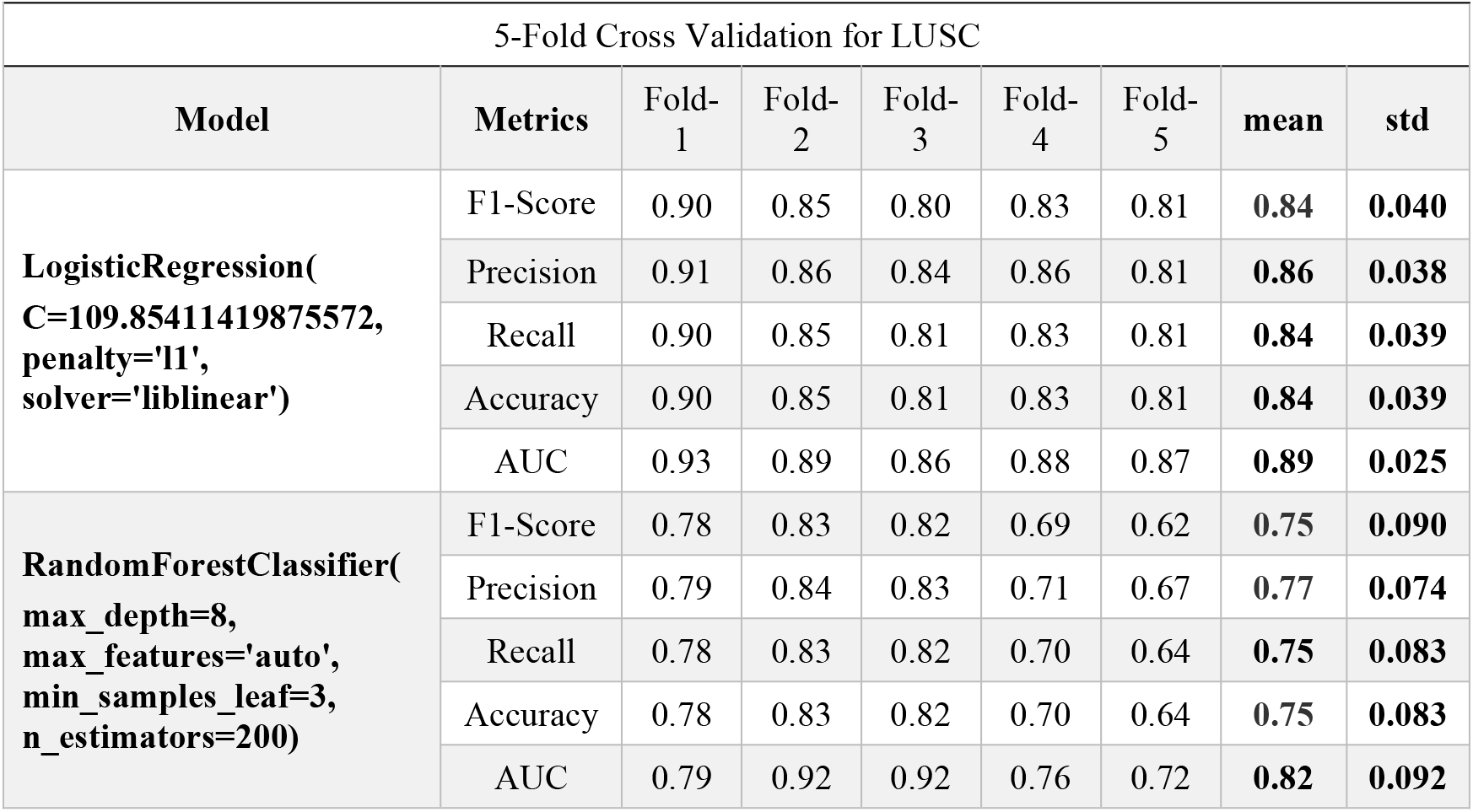
Accuracies of each fold for LUSC

**Table 6:**
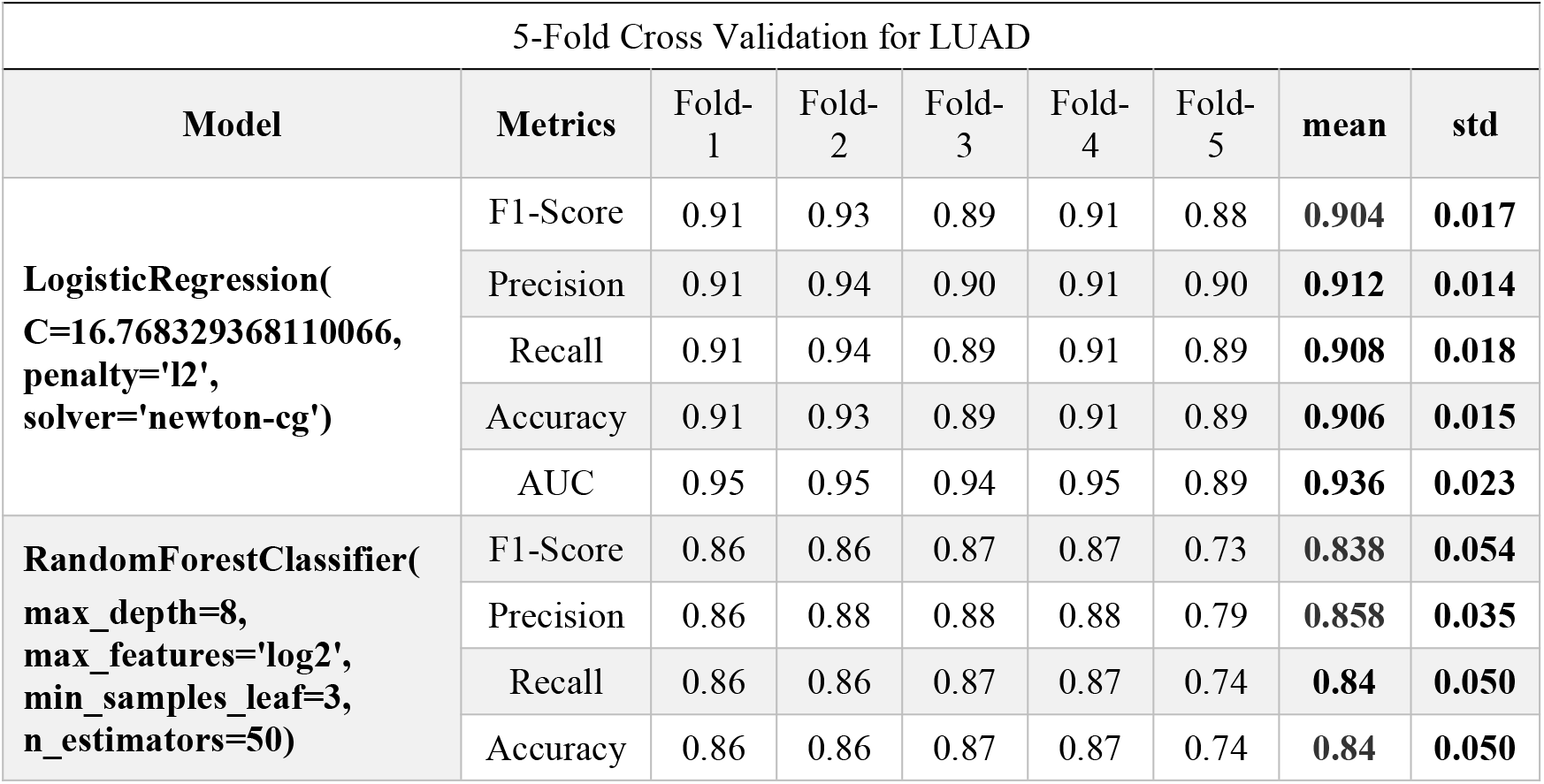
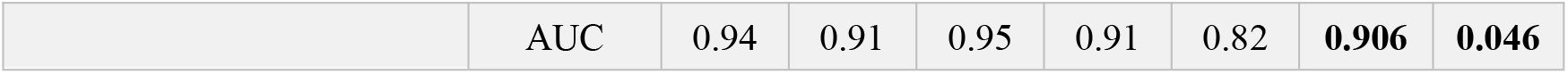
Accuracies of each fold for LUAD

### 4.2 Random Forest Feature Importance

The feature selection techniques select a subset of the most relevant features according to the target feature. The main goal of choosing the most relevant features is running the algorithms more efficiently to space and time complexity problems. Irrelevant input features can mislead the machine learning algorithms resulting in worse performance. In this work, we plotted the feature importances on two sets of features; 1) Full set of clinical features and 2) top 10 somatically mutated driver genes. Importance ranking of the features are provided by the fitted attribute feature_importances_of the scikit-learn Python machine learning library. The feature importances are computed as the mean and standard deviation of accumulation of the impurity decrease within each tree (Figure 6 and Figure 8). We also plotted the top 9 most correlated features to days_to_death (Figure 7 and Figure 9).

**Figure 6:**
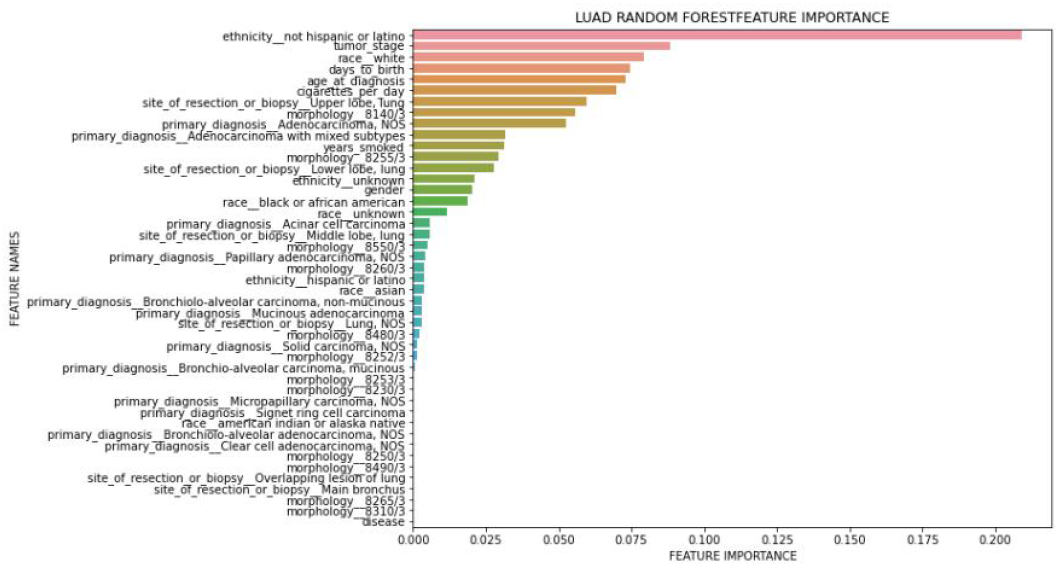
Importance ranks of LUAD clinical features only

**Figure 7:**
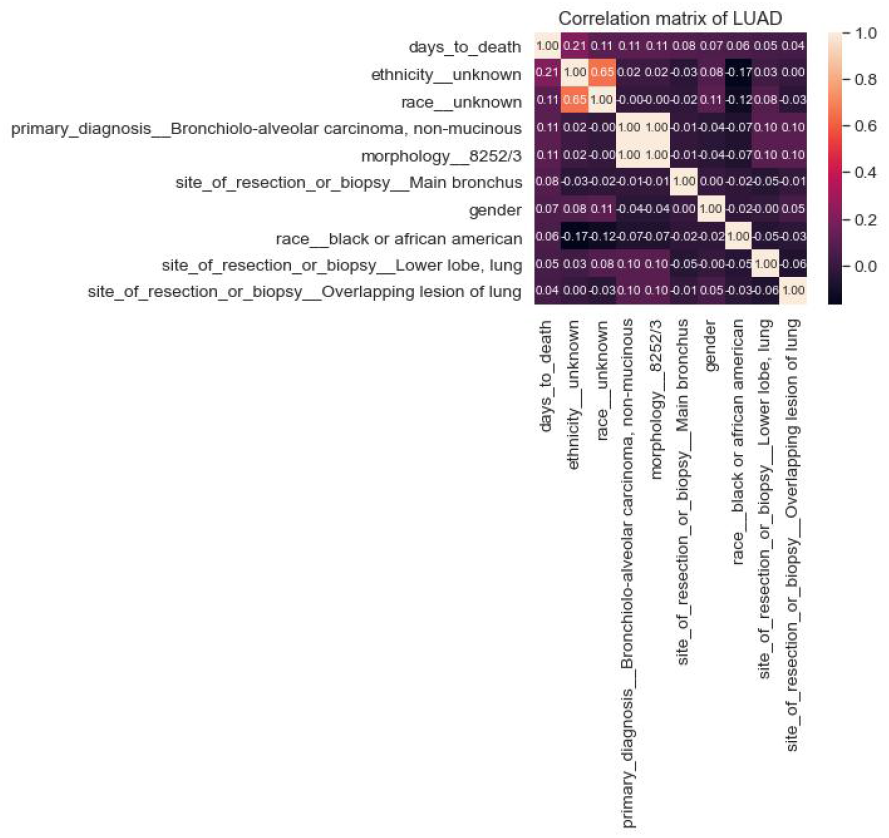
Top 9 most correlated LUAD clinical features to days_to_death

**Figure 8:**
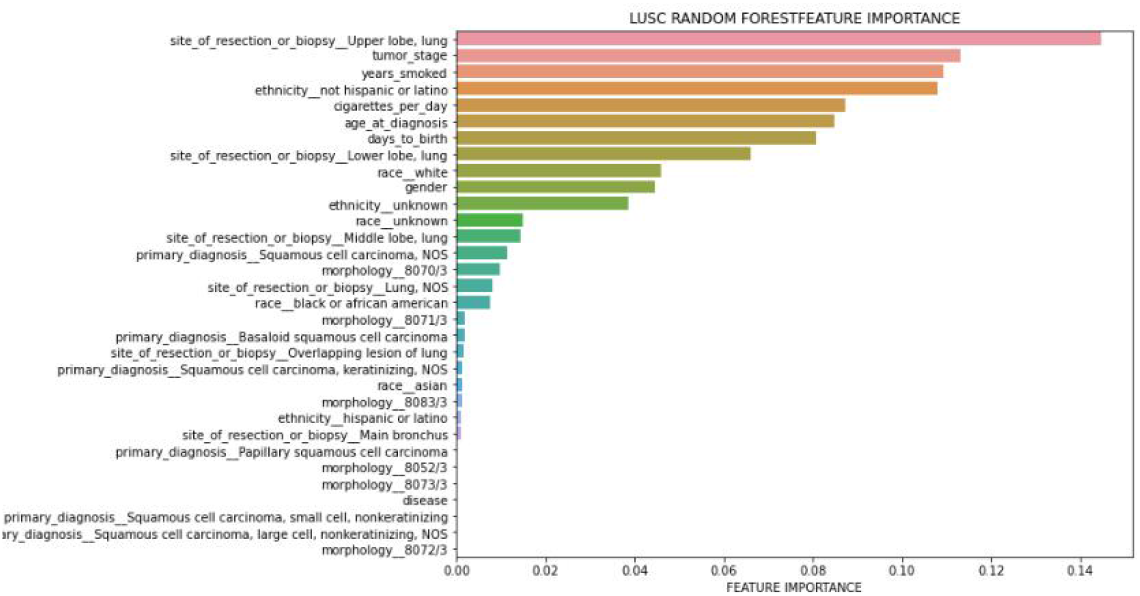
Importance ranks of LUSC clinical features only

**Figure 9:**
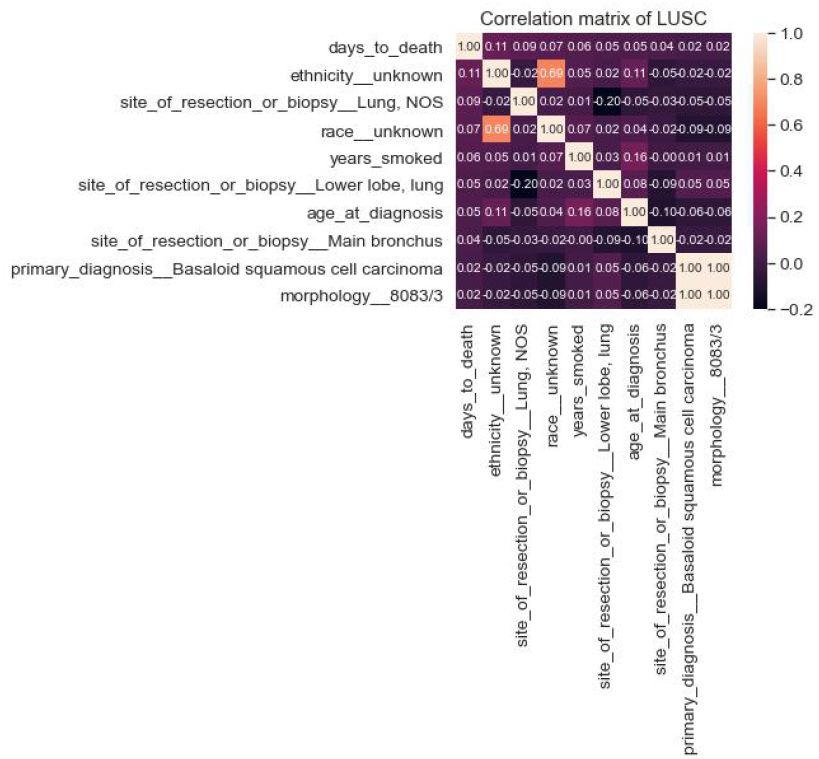
Top 9 most correlated LUSC clinical features to days_to_death

## 5. SOMATICALLY MUTATED GENES

In this section, we researched which somatically mutated genes effect the risk classification by feature engineering. For this, we combined the top 10 most highly mutated somatic driver genes as genomic features as well as the clinicopathological features. For this purpose, we selected the top 10 most highly mutated somatic driver genes for LUAD, and LUSC identified in our previous publication [16] using SominaClust[17].

The top 10 most somatically mutated driver gene features for LUAD patients were CDH10, COL11A1, CSMD3, HMCN1, KEAP1, KRAS, LRP1B, SPTA1 TP53, USH2A. When we tried to find a correlation between the risk of the patient (high-low) and these ten genes, KEAP1, TP53, USH2a and CSMD3 popped up in the top 10 feature list. We also observed that there is a coefficient correlation between ’KEAP1’ and risk of the patient. Nevertheless, there was no significant improvement in the performance of the LUAD Logistic Regression and Random Forest model after the addition of the mutated genes. (Figure 10 and Figure 11). Performance of each five-folds can be observed in Table 7 with a mean and standard deviation of folds. Moreover, KEAP1 mutation has higher feature importance followed by TP53, USH2A, CSMD3, LRP1B, SPTA1, CDH10, HMCN1, KRAS and COL11A1. Mutated genes have higher importance than many clinical features, however, ethnicity and race have highest feature importance, interestingly. Mostly used clinical variables such as age and tumor stage have higher importance than gene mutations. Site of resection, morphology, primary diagnosis, smoking amount have importance along with the gene mutations. Although adding gene mutations did not improve the performance, they have high importance with clinical variables, therefore they can be used together. For example, loss of function mutations in KEAP1 gene, promote KRAS-driven lung tumorigenesis [18] that may be reason of correlation of KEAP1 with risk of patients, therefore using KEAP1 and KRAS together with clinical variables can be considered.

**Table 7:**
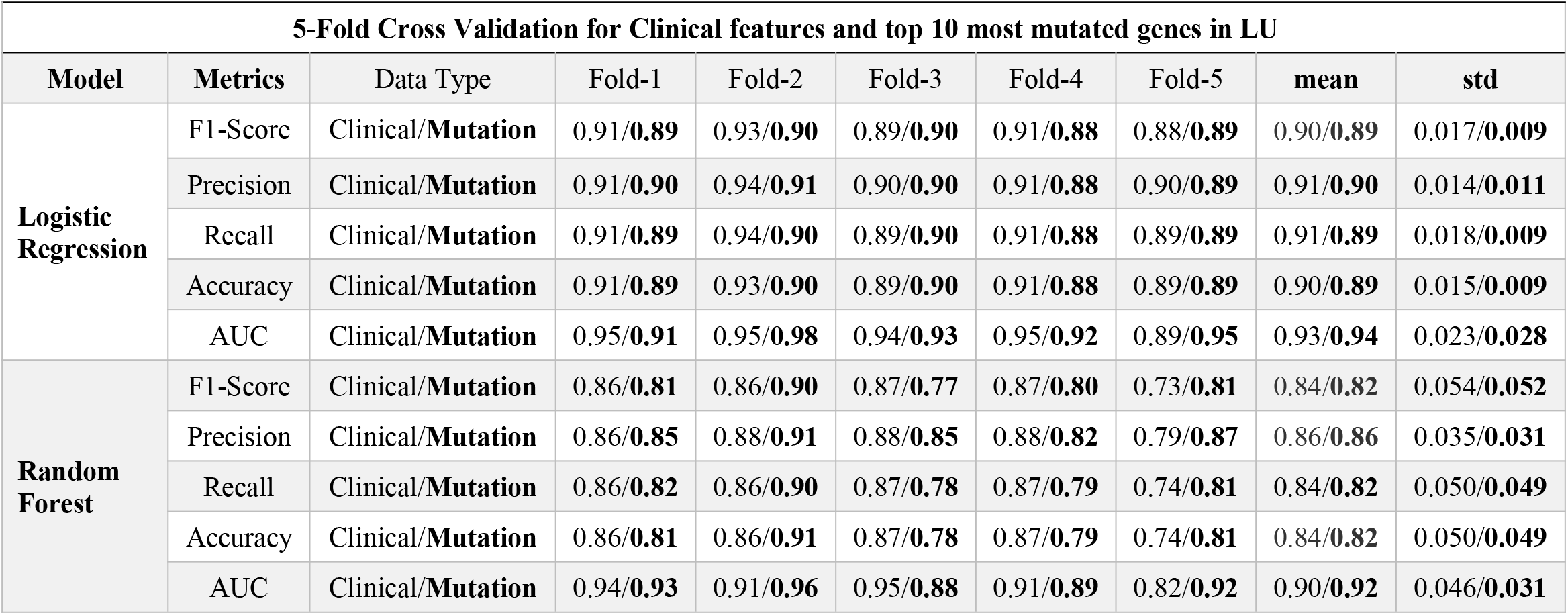
Accuracies of each fold for LUAD model with clinical features and top 10 somatically mutated genes

| 5-Fold Cross Validation for Clinical features and top 10 most mutated genes in LU |  |  |  |  |  |  |  |  |  |
| --- | --- | --- | --- | --- | --- | --- | --- | --- | --- |
| Model | Metrics | Data Type | Fold-1 | Fold-2 | Fold-3 | Fold-4 | Fold-5 | mean | std |
| Logistic Regression | F1-Score | Clinical/**Mutation** | 0.91/**0.89** | 0.93/**0.90** | 0.89/**0.90** | 0.91/**0.88** | 0.88/**0.89** | 0.90/**0.89** | 0.017/**0.009** |
|  | Precision | Clinical/**Mutation** | 0.91/**0.90** | 0.94/**0.91** | 0.90/**0.90** | 0.91/**0.88** | 0.90/**0.89** | 0.91/**0.90** | 0.014/**0.011** |
|  | Recall | Clinical/**Mutation** | 0.91/**0.89** | 0.94/**0.90** | 0.89/**0.90** | 0.91/**0.88** | 0.89/**0.89** | 0.91/**0.89** | 0.018/**0.009** |
|  | Accuracy | Clinical/**Mutation** | 0.91/**0.89** | 0.93/**0.90** | 0.89/**0.90** | 0.91/**0.88** | 0.89/**0.89** | 0.90/**0.89** | 0.015/**0.009** |
|  | AUC | Clinical/**Mutation** | 0.95/**0.91** | 0.95/**0.98** | 0.94/**0.93** | 0.95/**0.92** | 0.89/**0.95** | 0.93/**0.94** | 0.023/**0.028** |
| Random Forest | F1-Score | Clinical/**Mutation** | 0.86/**0.81** | 0.86/**0.90** | 0.87/**0.77** | 0.87/**0.80** | 0.73/**0.81** | 0.84/**0.82** | 0.054/**0.052** |
|  | Precision | Clinical/**Mutation** | 0.86/**0.85** | 0.88/**0.91** | 0.88/**0.85** | 0.88/**0.82** | 0.79/**0.87** | 0.86/**0.86** | 0.035/**0.031** |
|  | Recall | Clinical/**Mutation** | 0.86/**0.82** | 0.86/**0.90** | 0.87/**0.78** | 0.87/**0.79** | 0.74/**0.81** | 0.84/**0.82** | 0.050/**0.049** |
|  | Accuracy | Clinical/**Mutation** | 0.86/**0.81** | 0.86/**0.91** | 0.87/**0.78** | 0.87/**0.79** | 0.74/**0.81** | 0.84/**0.82** | 0.050/**0.049** |
|  | AUC | Clinical/**Mutation** | 0.94/**0.93** | 0.91/**0.96** | 0.95/**0.88** | 0.91/**0.89** | 0.82/**0.92** | 0.90/**0.92** | 0.046/**0.031** |

**Figure 10:**
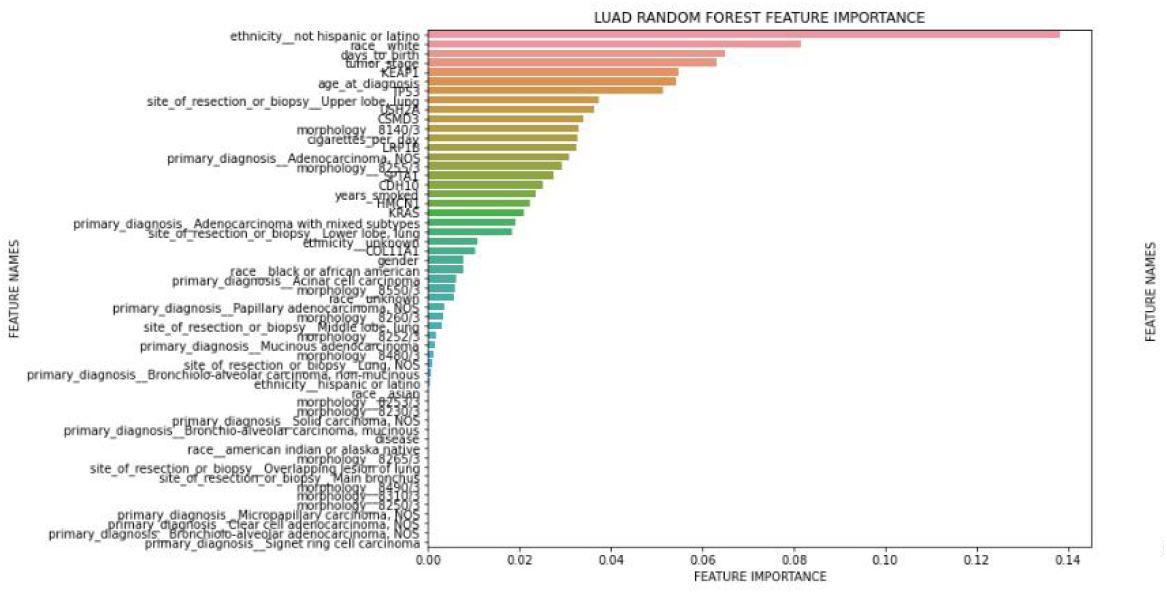
Importance ranking of LUAD top 10 somatically mutated genes and clinical features

**Figure 11:**
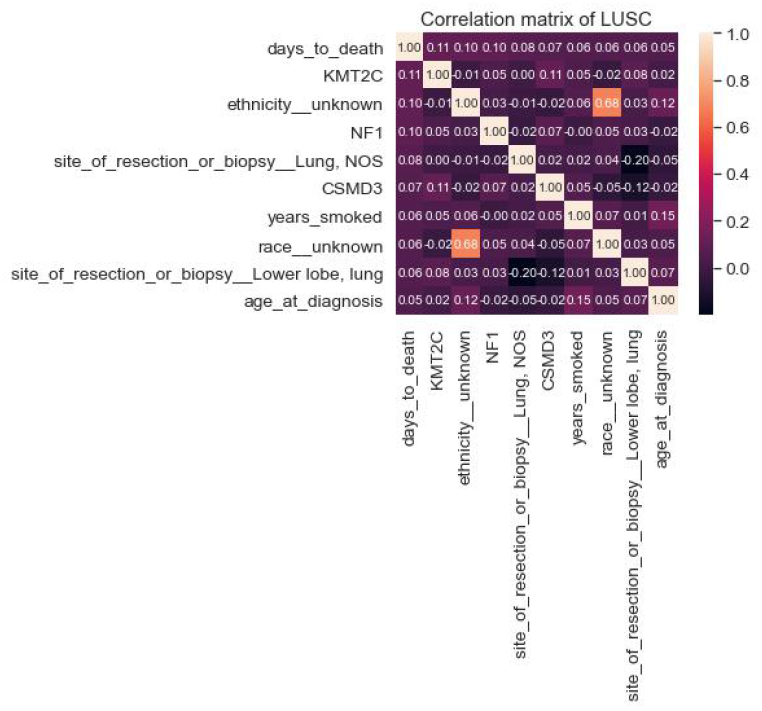
Top 9 most correlated LUAD clinical features and top10 somatically mutated genes to days_to_death

Although addition of the top 10 most highly mutated somatic driver genes to the classification model did not improve the performance of LUAD patients, they vastly improved the classification model of LUSC patients (Table 8). Top 10 somatically mutated genes of LUSC patients reported in our previous publication were CDKN2A, CSMD3, FAT1, KEAP1, KMT2C, KMT2D, NF1 NFE2L2, PIK3CA, TP53 [16]. The correlation between the risk of the patient (high vs low) and the given ten genes, the CSMD3 gene, impacted the model most by improving both the Logistic Regression and Random Forest performance in LUSC (Figure 12 and Figure 13). CSMD3 is one of the most frequently mutated genes in lung cancer and it is a potential tumor-suppressor [19]. Ethnicity is the second most important feature and smoking amount has high importance with age and tumor stage followed by TP53, KEAP1, NFE2L2, KMT2D, KMT2C, FAT1, CDKN2A, NF1 and PIK3CA gene mutations.

**Table 8:** Accuracies of each fold for LUSC model with clinical features and top 10 somatically mutated genes

| 5-Fold Cross Validation for Clinical features and top 10 most mutated genes |  |  |  |  |  |  |  |  |  |
| --- | --- | --- | --- | --- | --- | --- | --- | --- | --- |
| Model | Metrics | Data Type | Fold-1 | Fold-2 | Fold-3 | Fold-4 | Fold-5 | mean | std |
| Logistic Regression | F1-Score | Clinical/**Mutation** | 0.90/**0.89** | 0.85/**0.80** | 0.80/**0.87** | 0.83/**0.82** | 0.81/**0.85** | 0.84/**0.85** | 0.040/**0.036** |
|  | Precision | Clinical/**Mutation** | 0.91/**0.90** | 0.86/**0.82** | 0.84/**0.90** | 0.86/**0.83** | 0.81/**0.86** | 0.86/**0.87** | 0.038/**0.037** |
|  | Recall | Clinical/**Mutation** | 0.90/**0.89** | 0.85/**0.80** | 0.81/**0.87** | 0.83/**0.82** | 0.81/**0.85** | 0.84/**0.84** | 0.039/**0.036** |
|  | Accuracy | Clinical/**Mutation** | 0.90/**0.89** | 0.85/**0.80** | 0.81/**0.87** | 0.83/**0.83** | 0.81/**0.85** | 0.84/**0.85** | 0.039/**0.034** |
|  | AUC | Clinical/**Mutation** | 0.93/**0.93** | 0.89/**0.88** | 0.86/**0.93** | 0.88/**0.90** | 0.87/**0.91** | 0.89/**0.91** | 0.025/**0.021** |
| Random Forest | F1-Score | Clinical/**Mutation** | 0.78/**0.85** | 0.83/**0.75** | 0.82/**0.76** | 0.69/**0.81** | 0.62/**0.73** | 0.75/**0.78** | 0.090/**0.048** |
|  | Precision | Clinical/**Mutation** | 0.79/**0.87** | 0.84/**0.81** | 0.83/**0.80** | 0.71/**0.82** | 0.67/**0.78** | 0.77/**0.82** | 0.074/**0.032** |
|  | Recall | Clinical/**Mutation** | 0.78/**0.85** | 0.83/**0.76** | 0.82/**0.77** | 0.70/**0.81** | 0.64/**0.74** | 0.75/**0.79** | 0.083/**0.044** |
|  | Accuracy | Clinical/**Mutation** | 0.78/**0.85** | 0.83/**0.76** | 0.82/**0.77** | 0.70/**0.81** | 0.64/**0.74** | 0.75/**0.79** | 0.083/**0.044** |
|  | AUC | Clinical/**Mutation** | 0.79/**0.91** | 0.92/**0.86** | 0.92/**0.82** | 0.76/**0.88** | 0.72/**0.84** | 0.82/**0.86** | 0.092/**0.037** |

**Figure 12:**
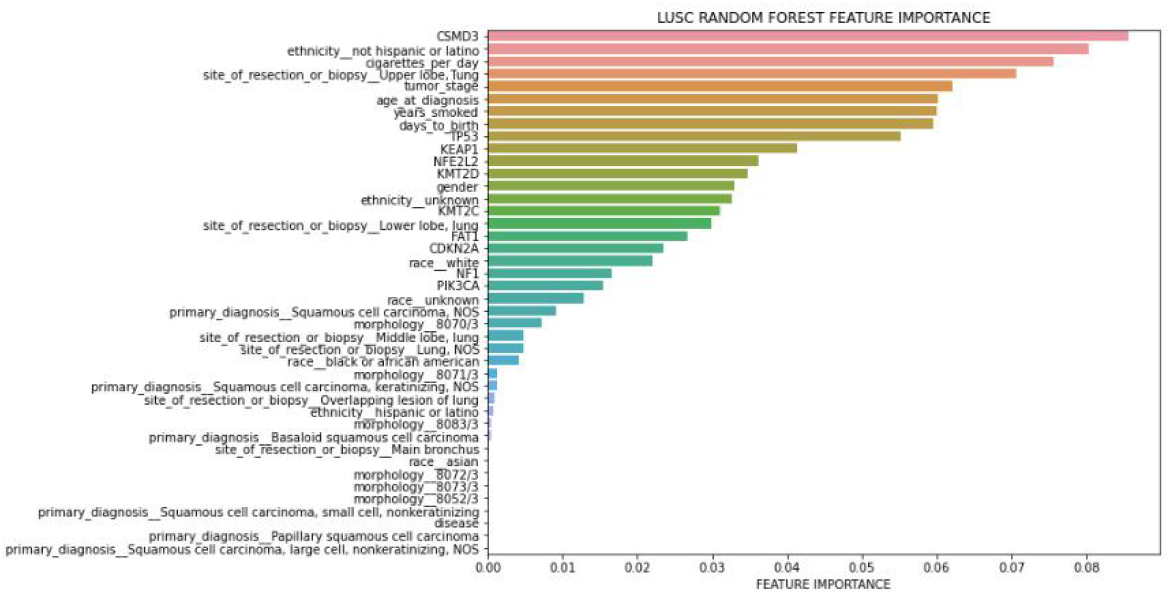
Importance ranking of LUSC top 10 somatically mutated genes and clinical features

**Figure 13:**
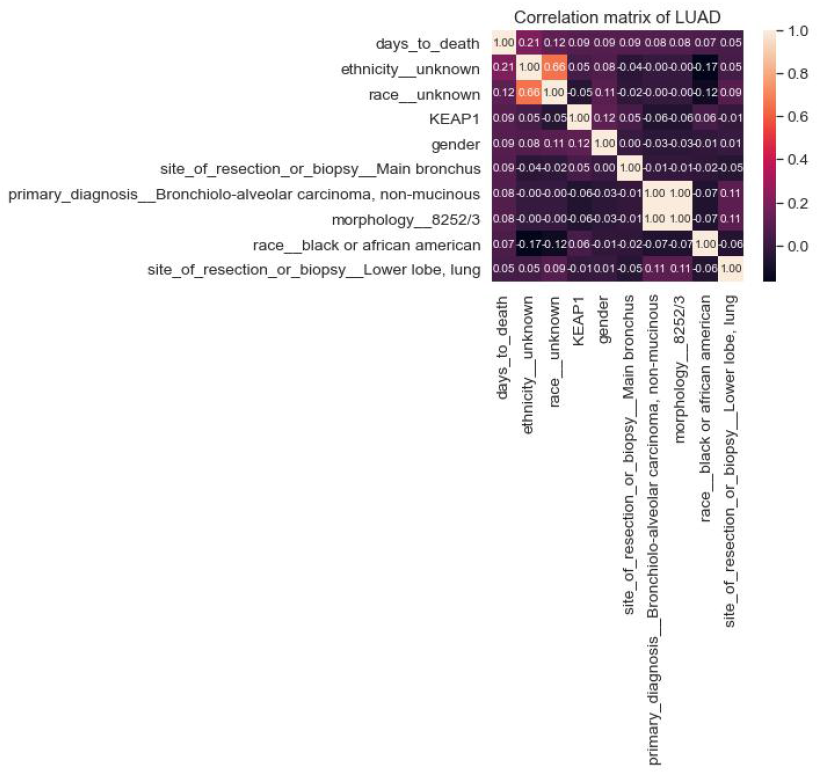
Top 9 most correlated LUSC clinical features and top10 somatically mutated genes to days_to_death

## 6 DISCUSSION

Main goal of our study was to investigate the clinical features or biomarker genes that are most helpful in prediction of risk stratification of lung adenocarcinoma and lung squamous carcinoma patients. For this purpose, we employed several machine learning studies investigating the vast clinical feature set and top 10 most somatically mutated gene set of TCGA lung adenocarcinoma and lung squamous carcinoma patients and ranked the features that contributed to the risk stratification most. Overall, clinic features still have importance even when gene mutations are added to analysis and can be considered to use with gene mutations. As a result of this analysis, new genes such as KEAP1 for LUAD and CSMD3 for LUSC with new clinicopathological features such as site of resection can be added to clinical decision processes. Future work of our model involves incorporating these developed machine learning models to a user-friendly web interface enabling both clinicians and lung cancer patients to assess the patients’ risk stratification.

## Data Availability

All data produced are available online at TCGA

